# Evaluating the Risk of Conflict on Recent Ebola Outbreaks in Guinea and the Democratic Republic of the Congo

**DOI:** 10.1101/2023.05.30.23290713

**Authors:** Gina E C Charnley, Nathan Green, Ilan Kelman, Espoir B Malembaka, Katy A M Gaythorpe

## Abstract

**Background:** Reducing Ebola virus transmission relies on the ability to identify cases and limit contact with infected bodily fluids through biosecurity, safe sex practices, safe burial and vaccination. Armed conflicts can complicate outbreak detection and interventions due to widespread disruption to governments and the populations. Guinea and the Democratic Republic of the Congo (DRC) have historically reported the largest and the most recent Ebola virus outbreaks. Understanding if conflict played a role in these outbreaks may help in identifying key risks factors to improve disease control.

**Methods:** We used data from a range of publicly available data sources for both Ebola virus cases and conflict events from 2018 to 2021 in Guinea and the DRC. We fitted these data to conditional logistic regression models using the Self-Controlled Case Series methodology to evaluate the magnitude in which conflict increased the risk of reported Ebola virus cases in terms of incidence rate ratio. We re-ran the analysis sub-nationally, by conflict sub-event type and tested any lagged effects.

**Results:** Conflict was significantly associated with an increased risk of reported Ebola virus cases in both the DRC and Guinea in recent outbreaks. The effect was of a similar magnitude at 1.88- and 1.98-times increased risk for the DRC and Guinea, respectively. The greatest effects (often higher than the national values) were found in many conflict prone areas and during protest/riot-related conflict events. Conflict was influential in terms of Ebola virus risk from 1 week following the event and remained important by 10 weeks.

**Conclusion:** Extra vigilance is needed following protests and riot-related conflict events in terms of Ebola virus transmission. These events are highly disruptive, in terms of access to transportation and healthcare and are often in urban areas with high population densities. Additional public health messaging around these types of conflict events, relating to the risks and clinical symptoms may be helpful in reducing transmission. Future work should aim to further understand and quantify conflict severity and intensity, to evaluate dose-response relationships in terms of disease risk.

## Background

Ebola is a virus belonging to the *Filoviridae* family and was first identified in 1976 in what are now the Democratic Republic of the Congo (DRC) and South Sudan [1,2]. Ebola virus disease (EVD) causes acute haemorrhagic fever, leading to mortality rates of ∼50%, in the absence of any specific treatment. The virus is spread through human-to-human transmission via contact with infected body fluids and fomites, with an average incubation period of 8 to 10 days [3,4].

Since its discovery, there have been several EVD outbreaks (with sporadic cases in Europe and North America) [5]. The worst recorded outbreaks, in terms of total cases and deaths, were the 2013-2016 West Africa outbreak, particularly in Sierra Leone, Liberia and Guinea and the 2018-2020 outbreak in the DRC. The most recent EVD outbreaks have been reported in Uganda (*Sudan Orthoebolavirus)*, Guinea (*Zaire Orthoebolavirus)* and the DRC (*Zaire Orthoebolavirus)*, all of which occurred in 2021 and 2022 [6].

EVD control is achieved through identifying cases and interventions which reduce the risk of contact with infected people, via biosecurity in EVD treatment centres, safe burial practices, reducing sexual transmission [7], timely detection (via strategically positioned decentralized laboratories) [8], contact tracing and post-mortem testing to identify EVD-related deaths, genomic sequencing (to infer spatiotemporal transmission dynamics) [9,10], and more recently with the rVSVΔG-ZEBOV-GP Ebola virus envelope glycoprotein vaccine and monoclonal antibody therapeutics [11,12]. Conflict can impact these control measures, reducing the capacity to identify and respond to outbreaks [13]. Furthermore, conflict can cause disruption to a range of basic services including transport, healthcare and water, sanitation and hygiene (WASH) [14].

Targeted attacks on healthcare centres and workers are not limited to specific diseases or geographic regions [15] and there have been several reported attacks on Ebola treatment centres, creating fear and avoidance in accessing them [16]. To protect staff and patients, security forces and military personnel are often required to carry out healthcare activities [10,15,17]. Additionally, fear and mistrust can lead to further attacks, sabotage and disruption of healthcare facilities and services. Distrust and dissatisfaction in the government, the military and international involvement via non-governmental organisations (NGOs) has also reduced the willingness to follow public health guidance and has increased vaccine hesitancy and misinformation [7,18].

In previous EVD outbreaks in the DRC and Guinea, conflict has been an ongoing issue to greater or lesser extents [19,20]. The DRC is currently experiencing armed conflict, particularly but not limited to the Kivu provinces. The conflict has been ongoing since the 1990s due to instability within the region, with many armed groups currently active along with governments’ involvement [21,22]. Guinea has experienced conflict, albeit to a lesser extent than in the DRC, mainly due to politically driven violence and demonstrations during post-election periods and excessive force by security forces [23].

Due to Guinea and the DRC having both the largest recorded outbreaks and the most recent EVD outbreaks [6], along with reported conflict, there is a need to further understand the mechanisms and evaluate the up-to-date conflict-related risks for recent EVD resurgences. Additionally, it is essential to understand the timing of conflict-related risks for EVD transmission and any lagged effects. Increasing our understanding of the magnitude and timing of risks can help EVD response planning and identify when extra surveillance may be needed.

Despite evidence showing possible links and effects of conflict on EVD, research to date is limited on magnitude, timings and mechanisms through which conflict impacts the risk of EVD, particularly qualitatively. Here, we use a methodology that has proved robust in evaluating the relationship between disease and conflict in previous studies [24] to address the following research aims:

1. Evaluate the risk of conflict on reported EVD cases during the 2018 to 2020 outbreak in the DRC and the 2021 outbreak in Guinea, both nationally and sub-nationally.
2. Evaluate if certain conflict types were more important than others in impacting EVD cases and suggest mechanisms for any differences found.
3. Examine the timing of conflict-related risk on EVD, including lagged effects.

## Methods

### Spatial Administrative Units

Subnational administrative boundaries differ between Guinea and the DRC, Table 1 presents each country’s definitions of subnational regions.

**Table 1.** Subnational administrative unit names for Guinea and the Democratic Republic of the Congo (DRC).

| Administrative Unit | Country | Name |
| --- | --- | --- |
| 1 | Guinea | Region |
| 1 | DRC | Province |
| 2 | Guinea | Prefecture |
| 2 | DRC | Territory |
| 3 | Guinea | Sub-prefecture |
| 3 | DRC | Sector |
| 4 | Guinea | Settlement |
| 4 | DRC | Settlement |

### Datasets

For the DRC, EVD data were collated from the DRC Ministry of Health (MoH) mailing lists [25,26] and from the Humanitarian Emergency Response Africa (HEMA) dataset [27]. The temporal scale of the data was daily and ranged from 04 April 2018 to 11 July 2020 for the MoH data and 06 February 2021 to 14 April 2021 for the HEMA data. The spatial scale for the DRC data was to administrative level 2 for the DRC MoH data and to administrative level 1 for the HEMA dataset (Table 1), including new reported case and death counts. For Guinea, the data ranged from 14 February 2021 to 19 June 2021 and included new and cumulative daily case counts by administrative level 1 (Table 1), extracted from the Government of Guinea situation reports [28]. For both the DRC and Guinea, temporal and spatial scales and ranges were chosen based on data availability and all data which were available were utilized for the analysis.

Conflict data were exported from the Armed Conflict Location & Event Data Project’s (ACLED, Wisconsin, United States) data export tool [29]. ACLED was chosen over other data sources (e.g., Uppsala Data Conflict Program [30]), as it contained more detailed categorisation of conflict event type, which were fundamental to the aims of this work. For each country, data were available from 1997 to the present, with each data entry point equating to a reported conflict. Conflicts were reported on a daily scale to longitude and latitude (in degrees), and by administrative level 1, 2, 3 and 4 (Table 1). The data were categorised by conflict event type (riots, protest, battles, explosions/remote violence, strategic developments and violence against civilians) and sub-event type and provided information on number of conflict fatalities (definitions of the event types, according to ACLED, are provided in Additional file 1).

Due to the granularity of the different datasets and for compatibility with the model (a single number was needed to define the time of each EVD case/conflict, see Model Structure and Fitting and Additional file 2), the temporal scale was set to continuous weeks from 01 January 2018 to 30 June 2021 (183 continuous weeks). Week number was not restarted at the beginning of the calendar year, to account for outbreaks/conflict which endured over a December to January period. Administrative level 1 for Guinea and administrative level 2 for the DRC were selected as the spatial granularity for model fitting (Table 1), due to data availability and discrepancies in finer scale location names among datasets. Weeks rather than continuous days were chosen, due to the potential for reporting lags in both the EVD and conflict datasets and due to the incubation period of Ebola virus (>1 week).

### Model Structure and Fitting

We used the Self-Controlled Case Series (SCCS) methodology which investigates the association between an exposure and an outcome event. The method aims to estimate the effect by comparing the relative incidence of the adverse events (EVD case) within an exposure period of hypothesised excess risk (conflict), compared to all other times (peace, according to the dataset used). The method is a case only method, meaning cases are the only subjects used to test the hypothesised effect. The method has the advantage of not needing separate controls (e.g., those not infected), by automatically controlling for fixed confounders that remain constant over the observational period. Cases are treated as individuals and then stratified in the modelling approach to control for the confounders [31]. The method uses conditional logistic regression (R function clogit(), package “survival” [32]) to estimate the effect of an exposure (conflict event) on an outcome event (EVD case, either suspected or confirmed), compared to all other times (peace) in an observation period (0-183 weeks). Maximum likelihood estimations were used, based on an assumed Poisson probability distribution [33,34]. All models were fit using R Studio with R version 4.3.0.

Both the exposure and event were transformed to binary outcomes, either being reported in the data in a specific week and administrative unit (1), or not (0), assuming that if a conflict or EVD case occurred, then it would have been correctly identified and reported. For conflict, the definition of a conflict event was the same for both countries. As the method is case-only, both a conflict event and EVD case had to be reported in the same administrative level (administrative unit 1 for Guinea and administrative unit 2 for the DRC, see Table 1) to be included.

To control for the fixed confounders, each exposure and event (in the same administrative unit) was allocated a unique number and used as a stratified term in the model. The event was then assigned a pre-exposure (start of observation period to exposure), exposure (exposure week) and post-exposure (exposure to end of observation period) period (see Additional file 2 for fitted data setup). The intervals between the three periods were logarithmically transformed and used as an offset term in the model. Offsetting the interval accounts for the possibility that a longer interval would result in a greater opportunity for the event (EVD case) to occur by chance, not due to its association with conflict (or peace).

The outcome event data consist of values *n_ijk_*, which take the values 1 or 0 corresponding respectively to whether an EVD case was reported or not, for individual *i* = 1, … *N*, in the *j*th week, *j* = 1, …, *J*, with conflict exposure *k* = 1,2, corresponding to peace or conflict. The length of time spent in a particular interval *ijk* is given by *e*_*ijk*_. The likelihood, *L*, is a function of *β_k_* which are the coefficients for conflict state *k*, and *α_j_* the coefficients for week *j*. The total likelihood is given by the following,

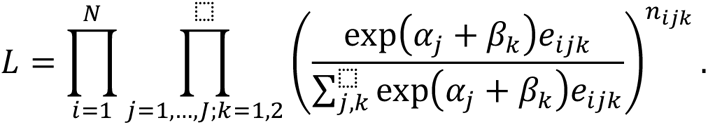

In standard survival analysis, *e*_*ijk*_ can be thought of as an offset term. Notice that non-time varying coefficients would drop-out of this self-controlled model. This is equivalent to a product multinomial likelihood. Hence, exp(*β*_2_ – *β*_1_) is the relative incidence for conflict, controlling for time (*β*_0_ = peace, based on the assumption that peace has no effect on EVD cases).

The model outcome variable was a log rate ratio, which was then exponentially transformed to incidence rate ratio (IRR), with a 95% confidence interval. IRR is the ratio of the incidence rates for individuals in a population during times of conflict exposure compared to times of peace (according to the dataset). The incidence rate is the rate at which incidence increases or decreases in a specific person-time (administrative level-week). IRR values of 1 indicate similar incidence rates in the conflict exposure period compared to the non-conflict exposure period, whereas an IRR > 1 suggests an increased risk and IRR < 1 indicates a decreased risk. Therefore, IRR quantifies the magnitude of the association in which conflict increased/decreased the risk of EVD transmission. IRR values were considered statistically significant at p < 0.05.

### By Sub-National Unit, Event Type and Lag

The model was fit several times to evaluate the different research questions above, either by sub-setting the data, or altering the way in which the exposure period was defined. The datasets were subset by the sub-national administrative level of the fitted data (see *Datasets* and Table 1) and the analysis repeated, to understand if certain geographic regions saw a greater magnitude of effect between Ebola and conflict. Similarly, conflict sub-event type (and conflict event type, results in Additional file 3) was also subset and the analysis repeated, to evaluate if certain types of conflict had a greater effect on EVD reported cases. Conflict event sub-type was chosen for the main analysis, rather than event type, as the event type names are generally more ambiguous and less descriptive, making the interpretation of the results more challenging.

The impact of lag was explored, based on the assumption that the effect of a conflict event on EVD cases may not be immediate (EVD’s incubation period is 8-10 days [3]) and may be long-lasting. Different exposure periods were tested, extending the period from 1 week following the conflict to 2, 4, 6, 8 and 10 weeks following the exposure, moving beyond previous work on EVD that tested the effect of conflict to 4 weeks [18]. However, a longer exposure period may result in a diluting effect due to a greater chance of an event (EVD case) being reported in an exposure period (conflict). To account for this, we tested the effect of a single week, 2, 4, 6, 8 and 10 weeks following the week the exposure was reported, rather than extending the exposure period.

### Quantifying conflict severity and proximity

To explore mechanisms through which the most impactful types of conflict (battles, protests and riots) may impact EVD transmission, we quantified conflict severity using two metrics. First, we presented conflict fatalities as a metric of severity (higher mortality = more severe conflict), both in terms of raw numbers for each country and conflict event type and as a proportion of total conflict fatalities. Second, we measured the proximity of all battles, protests and riots to major cities by the shortest distance between the coordinates of the conflict and the city (measured in longitude and latitude as degrees) in both the DRC [35] and Guinea [36]. We then compared the difference between the average distance for each conflict event type, to the total average distance for all event types, over the observation period of 2018 to 2022. The distance was measured using the Haversine method, which assumes a spherical earth, ignoring ellipsoidal effects (R functions distHaversine() and distm(), package “geosphere” [37]). We hypothesised that conflict types which more often occurred near more people (e.g., closer to major cities), would be more severe, and therefore have a bigger effect on disease transmission.

## Results

### Datasets

During the observation period, there were 12,237 EVD cases reported in 94 weeks (out of 183 weeks in the observation period) across 14 administrative 2 units for the DRC (all in Nord-Kivu, Sud-Kivu, Ituri and Équateur). For Guinea, there were 18 weeks (out of 183) which reported 1,797 EVD cases in two administrative 1 units (N’Zérékoré and Conakry). These cases corresponded to 168 (DRC) and 86 (Guinea) weeks of reported conflict exposure in the dataset during the observation period for the same corresponding administrative levels as the reported EVD cases. In the DRC, most EVD cases were reported in late 2018 to late 2019, while for Guinea, most cases were reported in 2021 (see Fig. 1).

**Fig. 1.**
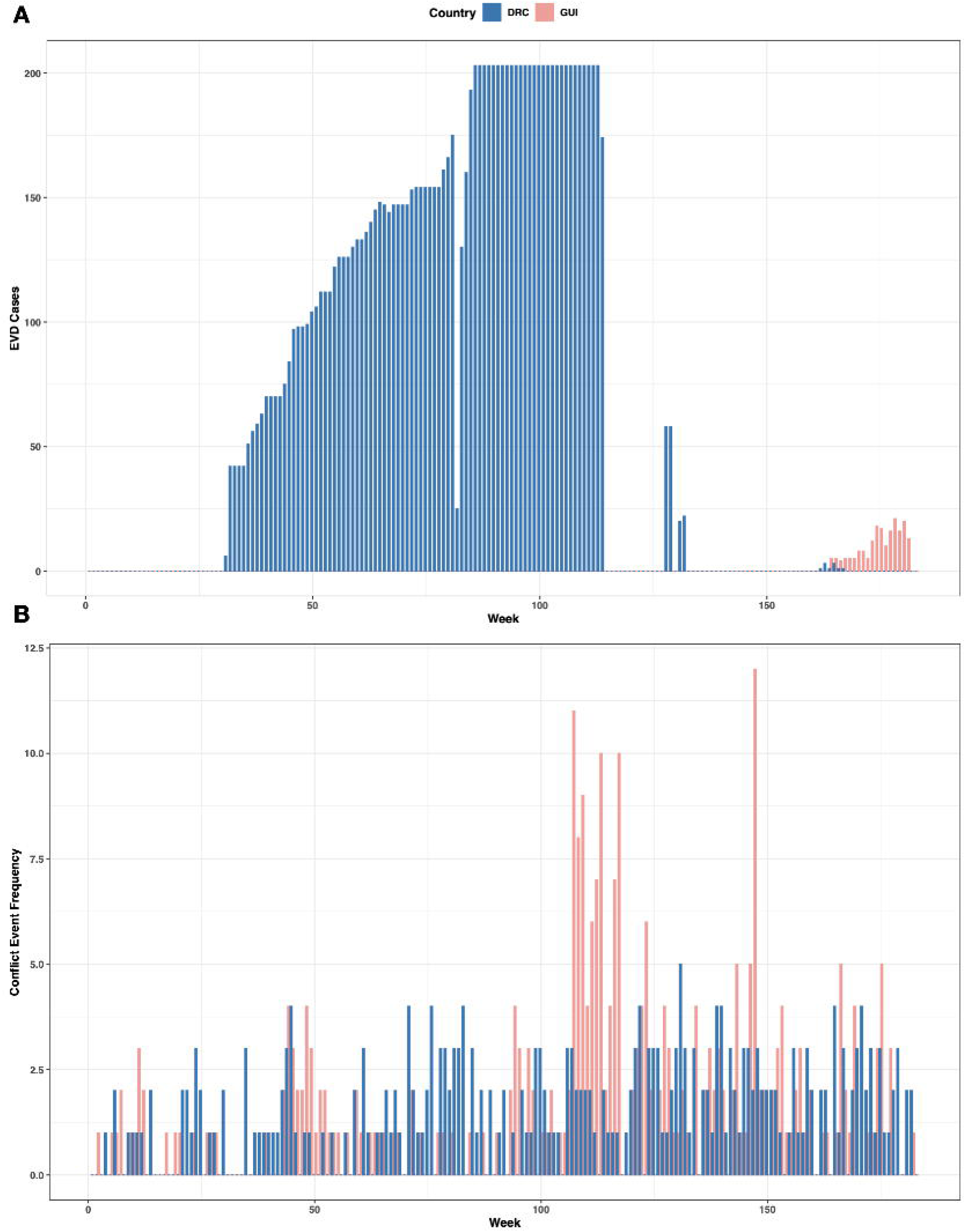
Weeks which reported **A,** Ebola virus disease (EVD) cases and **B,** conflict exposure for the Democratic Republic of the Congo (DRC) and Guinea (GUI).

In the DRC, EVD cases for the 2018-2020 outbreak were commonly reported in areas of high conflict frequency, namely the Kivu provinces. Similar results were found for Guinea, with all EVD cases being reported in the two regions that reported the highest conflict event frequency (Conakry and N’Zérékoré), but with overall lower levels of reported conflict compared to the DRC. The most frequent conflict sub-event types (event types) in the fitted data varied by country. For the DRC, these included armed clashes (battles), attacks (violence against civilians) and peaceful protests (protests). In Guinea, the most reported conflict sub-event types (event types) were violent demonstrations (riots), peaceful protests (protests) and mob violence (riots) (Table 2).

**Table 2.**
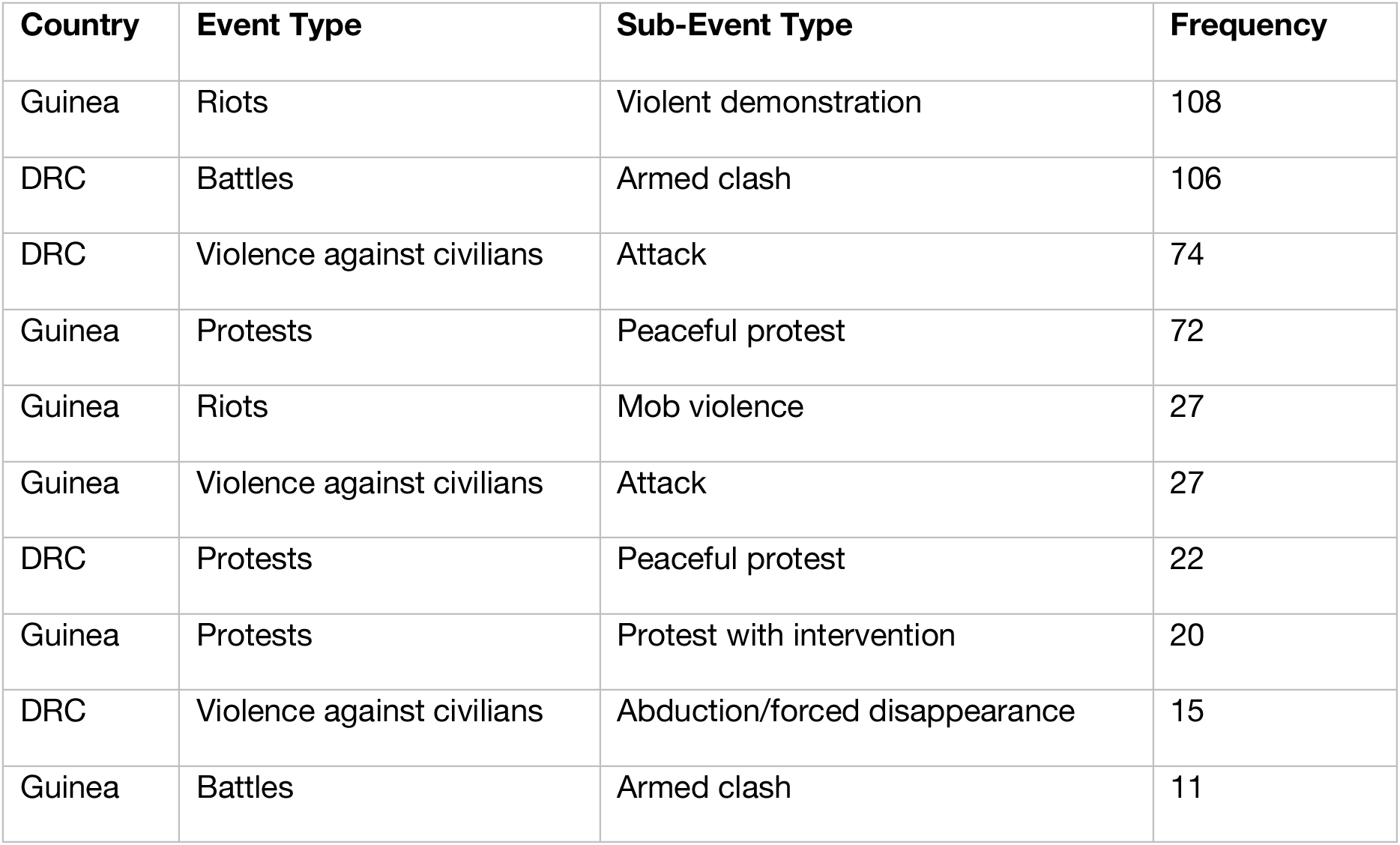
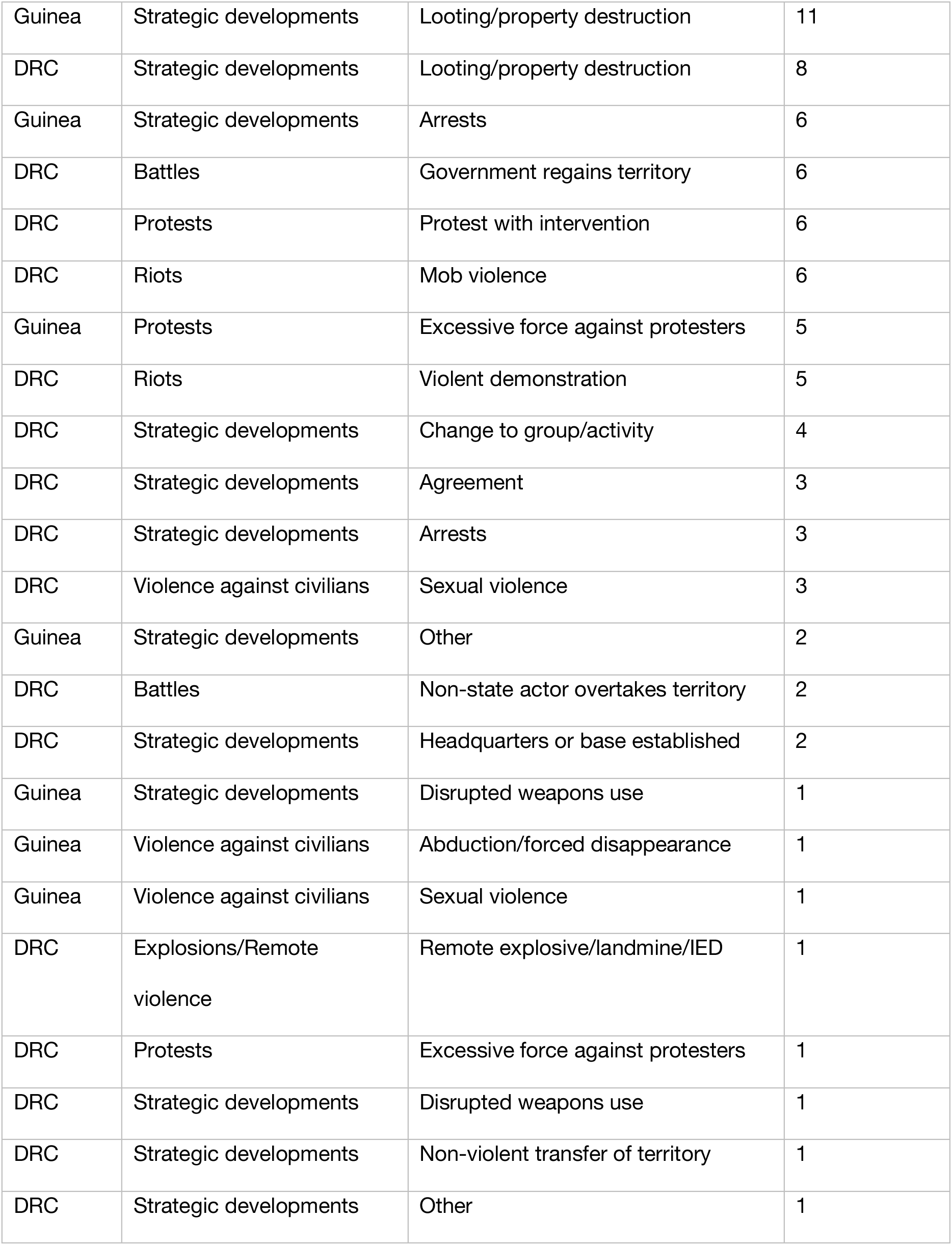
Frequency of conflict event types & sub-event types in the data fitted to the model for the Democratic Republic of the Congo (DRC) and Guinea.

### Model Output

Conflict was significantly associated with an increased risk of reported EVD cases in both the DRC and Guinea between 2018 and 2021. The magnitude of effect was similar for both countries, with conflict increasing the risk of EVD by 1.88 (1.76-2.02 95%Confidence Interval (CI)) times in the DRC, and 1.98 (1.11-3.51 95%CI) times in Guinea.

### By Sub-National Unit

The effect of conflict on EVD varied sub-nationally in terms of magnitude and uncertainty (Fig. 2). In the first week of the reported conflict exposure, the administrative units with the highest reported effect were Mbandaka (Équateur, DRC), Mwenga (Sud-Kivu, DRC), Goma (Nord-Kivu, DRC), Butembo (Nord-Kivu, DRC) and N’Zérékoré (Guinea) with IRR values of 4.26 (0.58-31.2 95%CI), 2.49 (1.87-3.31 95%CI), 2.19 (1.69-2.83 95%CI), 2.12 (1.76-2.57 95%CI) and 2.06 (1.13-3.77 95%CI), respectively. Some areas did not present significant results, often having higher uncertainty, including Aru, Bolomba, Ingende, Mbandaka in the DRC and Conakry in Guinea.

**Fig. 2.**
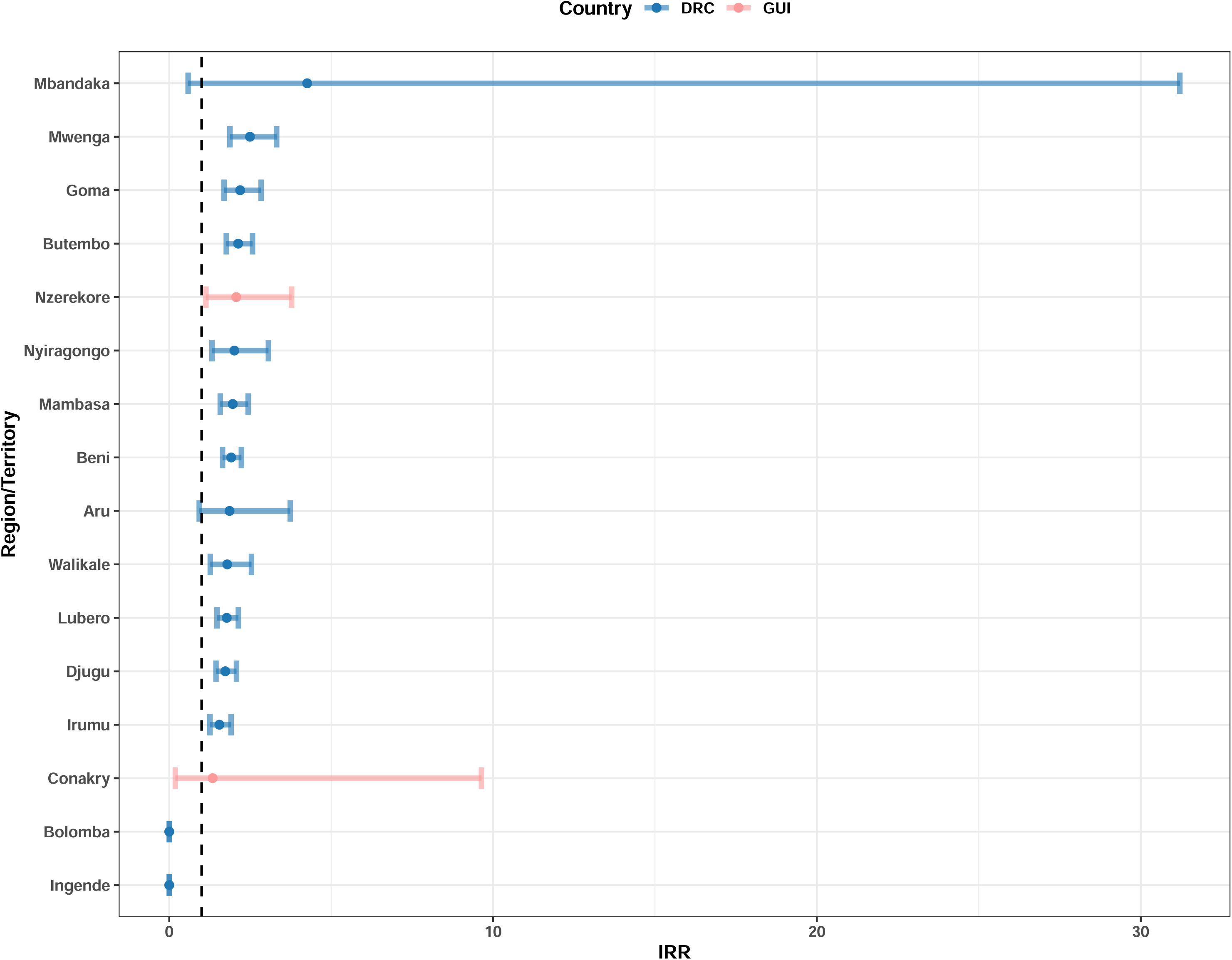
Incidence rate ratio (IRR) for the effect of conflict event exposure on EVD cases for the Democratic Republic of the Congo (DRC) and Guinea (GUI) during the week the conflict was reported at sub-national administrative unit.

### By Event Type

The greatest impact for conflict event sub-type was armed clashes in Guinea (IRR = 3.45, 0.80-14.8 95%CI) (Fig. 3); however, this was not statistically significant, along with 12 other conflict sub-types in Guinea (abduction/forced disappearances, arrests, attacks, change to group/activity, disrupted weapons use, excessive force against protesters, looting/property destruction, mob violence, other, peaceful protests, protests with intervention and sexual violence). Violent demonstrations had the greatest significant impact on EVD cases in Guinea causing a 2.38 (1.04-5.41 95%CI) times increased risk of EVD.

**Fig. 3.**
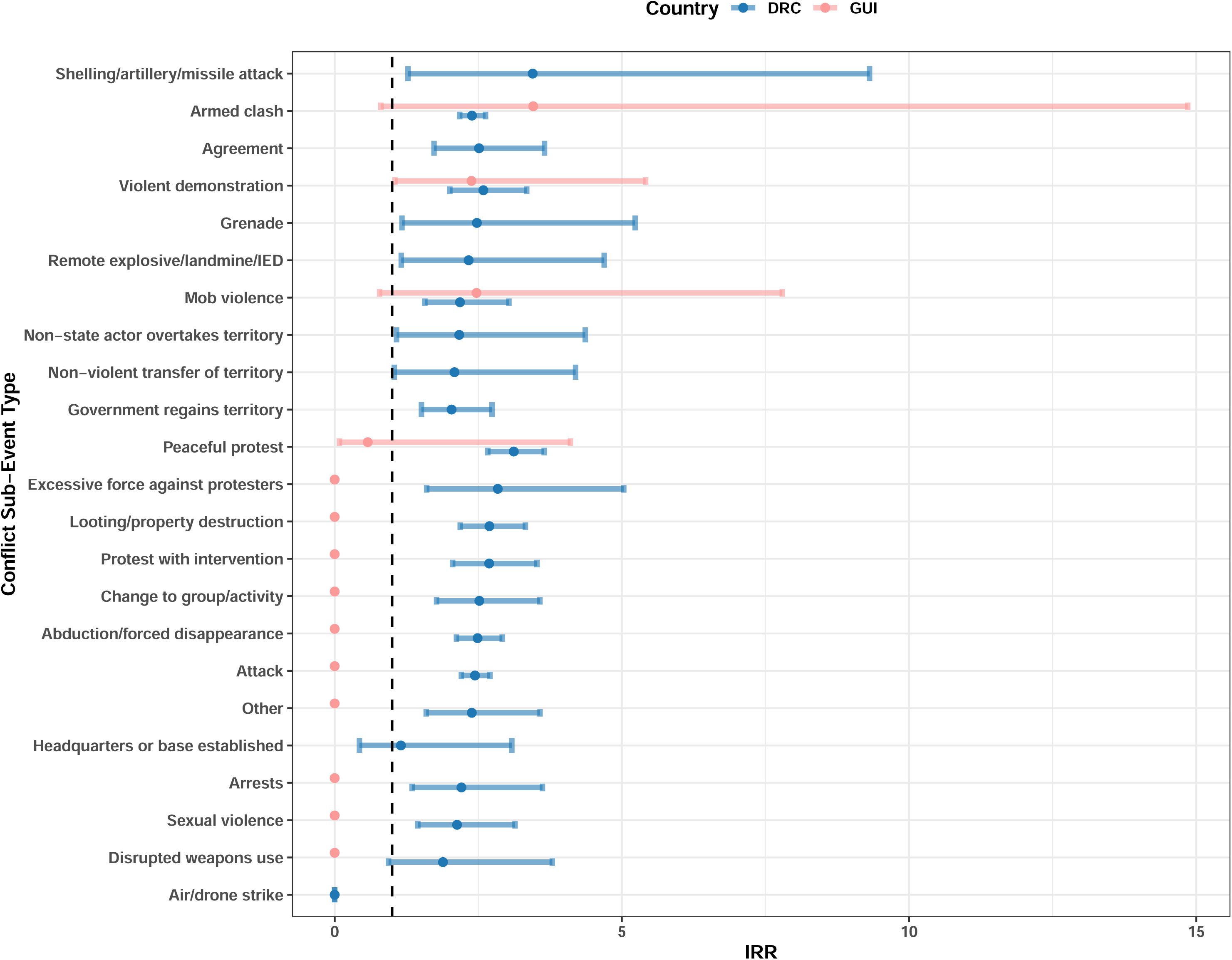
Incidence rate ratio (IRR) for the effect of conflict event exposure on EVD cases for the Democratic Republic of the Congo (DRC) and Guinea (GUI) during the week the conflict was reported by conflict sub-event type.

Only three conflict event sub-types were not significantly associated with EVD incidence in the DRC: air/drone strikes, disrupted weapons use and headquarters or base establishments (mainly due to very small sample sizes). For the conflict sub-types that did have a significant effect, shelling/artillery/missile attack had the biggest effect on EVD cases at 3.44 (1.27-9.3195%CI) times increased risk, followed by peaceful protests (IRR = 3.11, 2.66-3.64 95%CI), excessive force against protesters (2.83, 1.60-5.03 95%CI), looting/property destruction (2.69, 2.18-3.31 95%CI), protests with interventions (2.68, 2.05-3.52 95%CI) and violent demonstrations (2.58, 2.00-3.34 95%CI) (Fig. 3). For the results regarding conflict event type (rather than sub-event types), see Additional file 3.

### By Lag

All lag periods were found to be statistically significant for both countries. Extending the lag periods caused the effect of conflict on EVD cases to be almost eliminated (IRR = 1) by 10 weeks in the DRC, with an IRR of 1.05 (1.02-1.09 95%CI). In Guinea, the risk of EVD cases decreased to 1.41 (0.99-2.01 95%CI) times at 4 weeks and then increased to 1.72 (1.38-2.14, 95%CI) by 10 weeks. The uncertainty was far greater in Guinea with slightly less significant results (according to p values) compared to the DRC (see Fig. 4).

**Fig. 4.**
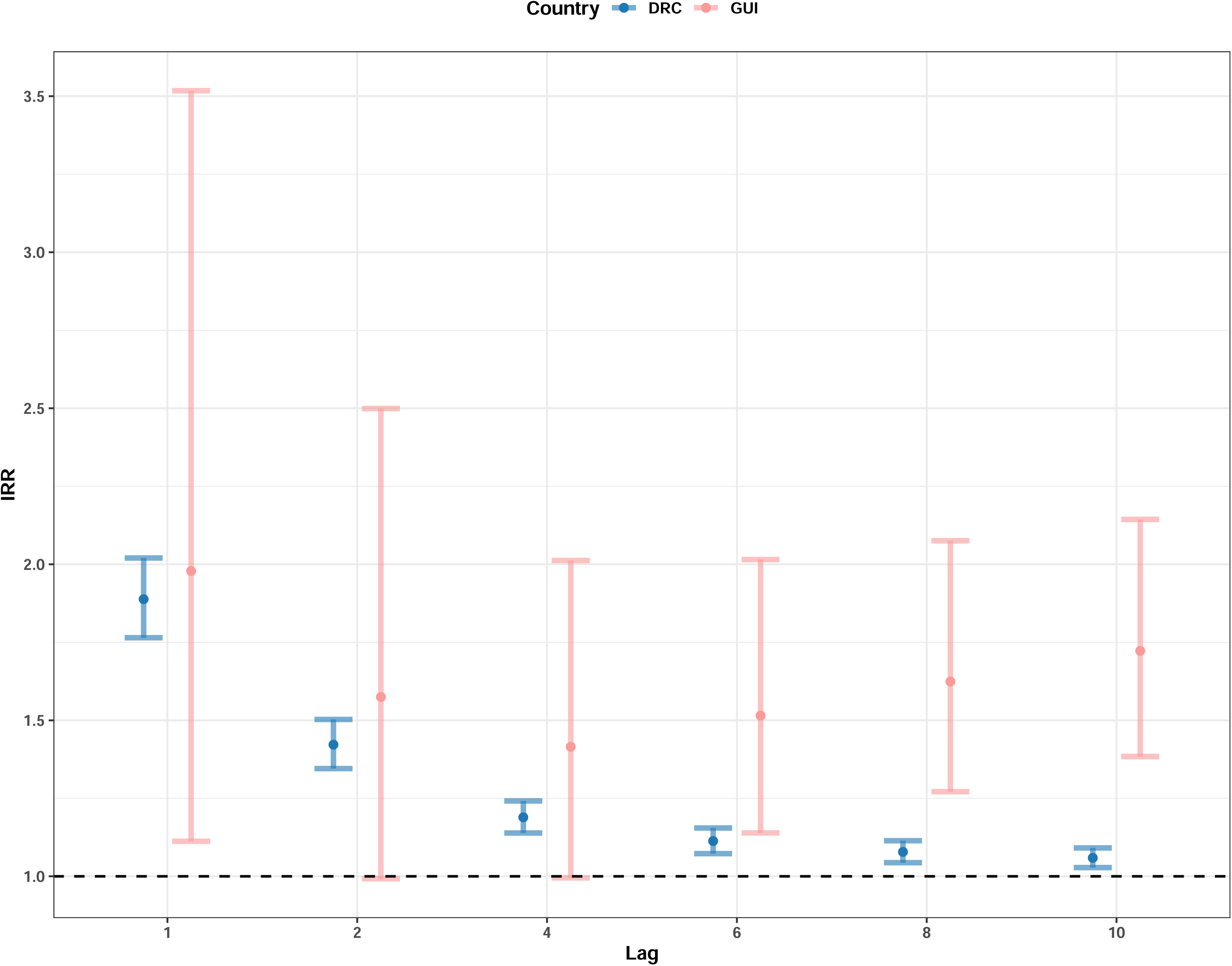
Incidence rate ratio (IRR) for the effect of conflict event exposure on EVD cases for the Democratic Republic of the Congo (DRC) and Guinea (GUI) from the week the conflict was reported to 10 weeks following the exposure, by extending the exposure periods.

The lagged exposure periods for a single week 2, 4, 6, 8 and 10 weeks following the exposure resulted in conflict having a greater and more statistically significant (according to p values) impact on EVD cases (Fig. 5), compared to extending the periods (from 1 week to 10 weeks). All lag periods saw an increased risk from week 1 to week 10, with Guinea having a greater effect (although with wider ranging uncertainty), potentially due to the increase already found in Guinea at week 4 with the extended exposure periods (Fig. 4). The risk of EVD cases increased from 1.88 (1.76-2.02 95%CI) to 1.96 (1.83-2.10 95%CI) times in the DRC and from 1.96 (1.11-3.51 95%CI) to 3.30 (2.12-5.13 95%CI) times in Guinea by the 10th week following the event.

**Fig. 5.**
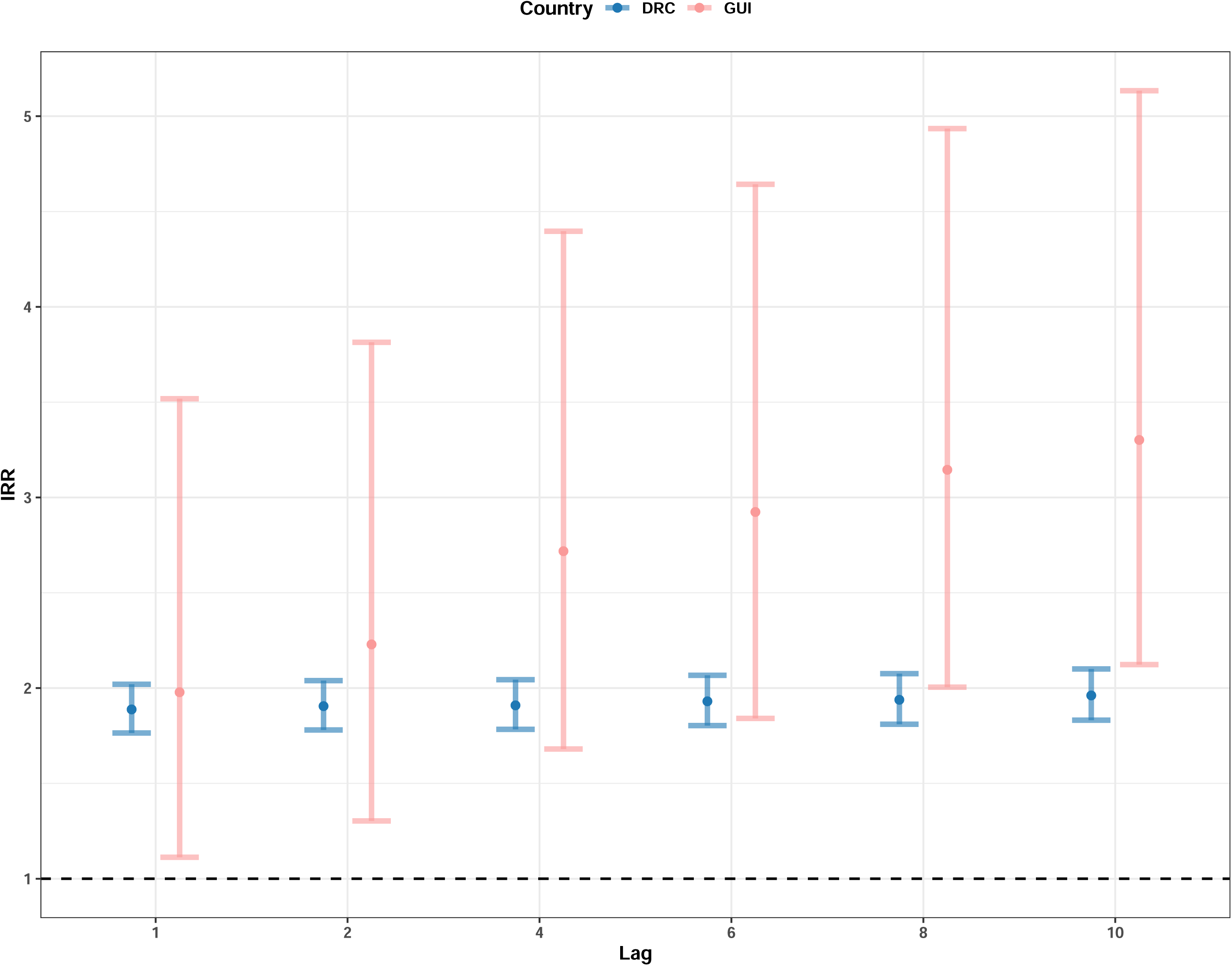
Incidence rate ratio (IRR) for the effect of conflict event exposure on EVD cases for the Democratic Republic of the Congo (DRC) and Guinea (GUI) from the week the conflict was reported to the 10th week following the conflict report, as a single week.

### Quantifying conflict severity and proximity

Table 3 shows the full results for the conflict fatalities and proximity analysis for each of the three most significant event types (battles, riots and protests). In the DRC, battles caused 56% of all conflict fatalities in the fitted data here (n = 7,900), compared to 0.19% (n = 27) and 3.06% (n = 430) for protests and riots, respectively. In Guinea, 64% (n = 189) of conflict fatalities were caused by riots, followed by 17.2% (n = 51) in battles and 7.12% (n = 21) in protests.

**Table 3.** Battles, riots and protests in terms of EVD risks by number of conflict fatalities, the percentage of total fatalities and the difference in average proximity from conflict events to major settlements (event type average distance minus all conflicts average distance) for the full four-year observation period for each country.

| Country | Event Type | Fatalities Frequency | Fatality Percentage | Difference in proximity |
| --- | --- | --- | --- | --- |
| DRC | Battles | 7,900 | 56.2% | 6.33km |
| DRC | Riots | 430 | 3.05% | -3.33km |
| DRC | Protests | 27 | 0.19% | -2.99km |
| Guinea | Battles | 51 | 17.3% | 17.2km |
| Guinea | Riots | 189 | 64.1% | -5.73km |
| Guinea | Protests | 21 | 7.11% | -11.4km |

On average, over the four-year period, battles were further away from major cities in both countries, followed by protests and riots in the DRC and riots and then protests in Guinea (for maps, see Additional file 4: Fig. S2). Compared to the average distance between all conflict events and major settlements, battles were 6.33km further away in the DRC and 17.2km further away in Guinea. In the DRC, protests were 2.99km closer and riots 3.33km closer, compared to the average distance. For Guinea, protests and riots were 11.4km and 5.73km closer, respectively.

## Discussion

During recent EVD outbreaks in 2018 to 2021, we quantified the risk of conflict event exposure and found an increased risk at a national level by a similar magnitude in the DRC and Guinea, at 1.88 and 1.98 times, respectively. The uncertainty in the IRR values for Guinea throughout all the analyses was much wider than the DRC. In the distribution of the exposures and events data fit to the model, there was overlap with areas of high reported conflict and EVD case frequency. Several of the regions/territories analysed had higher IRR values than the national risk: Mbandaka, Mwenga, Goma, Butembo (all in the DRC) and N’Zérékoré (in Guinea), which ranged from 2.06 to 4.26.

We evaluated if certain types of conflict may be more impactful than others, to contribute to our understanding of how conflict can change disease risk. The conflict sub-event types with the greatest impacts in terms of EVD risk were armed clashes in Guinea and shelling/artillery/missile attack in the DRC. However, both armed clashes and shelling/artillery/missile attacks had the largest uncertainty. Violent demonstrations had the strongest and most significant impact in Guinea, while in the DRC, peaceful protests, excessive force against protesters, looting/property destruction, protests with interventions and violent demonstrations had the largest, most significant impact, with little uncertainty.

The cornerstone of EVD control is through identifying cases and reducing contact with infected bodily fluids (and vaccination) [8,10]. Conflict can impact these control measures in multiple ways. In previous EVD outbreaks, weak health systems (due to access, supplies, safety of staff and patients and fear), complex socio-cultural and political environments (creating mistrust in governments and NGOs) and ineffective messaging and community engagement have all created problems in EVD response [20,38]. Additionally, there have been difficulties in administering EVD vaccines due to damaged transportation and infrastructure, although continued efforts aim to reach those most at risk [11,16,39]. Conflict and EVD can create a positive feedback loop, and it has been previously observed that conflict increases more during EVD outbreaks compared to controls [18,19]. A suggested explanation is that EVD outbreaks may further destabilise fragile regions and fuel conflicts. For example, mistrust increased in the DRC during the 2018-2019 EVD outbreak due to the government’s decision to ban populations in the affected provinces from participating in the presidential election [18,19].

A potential explanation for the large number of protest-related impacts on EVD cases found here, may be due to these events being more disruptive to a greater number of people. Protests rely on large congregations of people, increasing contact among individuals and therefore fueling EVD transmission. Furthermore, protests and demonstrations may be more likely to impact transportation, education and healthcare, due to the widespread disruption they can cause in dense urban areas [40,41]. Disruption of healthcare and transportation may be particularly damaging in controlling EVD, as it could complicate contact tracing and delay testing, diagnosis and treatment. Further evidence for this explanation may be that within these very impactful sub-events, looting/property destruction was also found highly impactful and important, which is often the product of protests and demonstrations and results in damage to healthcare infrastructure and supplies.

During 2018 to 2021, battles were the furthest away from major settlements in both countries, compared to protests and riots. Proximity may impact the effect of the conflict on the population, both in terms of the number of people affected and the magnitude of the effect. Proximity may also add further evidence to the above hypothesis that protests and demonstrations, although potentially less violent, may be more impactful than battles and armed clashes. Battles (in terms of the ACLED’s definition [29]) are more likely to be fought on pre-defined battle “grounds”, which are less likely to be close to settlements. Alternatively, these settlements, including their healthcare and testing facilities are quickly displaced and abandoned following the onset of conflict due to the immediate threat to life [42]. The absence of testing facilities may result in cases not being identified and reported, therefore making battles appear less impactful to EVD transmission.

A potential explanation for why remote violence/explosions and battles were very impactful but often with higher uncertainty is that these types of conflict occurred less frequently, but when they did, they had a very large impact on EVD cases. The fewer events therefore resulted in the large uncertainty and more data (if available) may help to understand if the larger impact of these conflict types is consistent through time. Additionally, peoples’ behaviour and risks must be considered during more “extreme” forms of violence such as battles and explosions, compared to protests and demonstrations. Violent conflict may have a very large initial impact (accounting for the high IRR values), but then the risk stabilises in the area as civilians take precautions (e.g., displacement) to protect their life [43].

Alternatively, there is the potential for more violent forms of conflict (such as battles and explosions) to decrease the risk of EVD. People may be more likely to stay at home during these events or be subject to curfews and not mix in public spaces, decreasing their exposure to the pathogen [44]. Furthermore, severe conflict may be more fatal (of both civilians and combatants), reducing the susceptible pool for EVD cases. More work is needed to understand how to quantity conflict severity for scientific research purposes, which could be used to identify dose-response relationships with disease, while still considering that conflicts impact individuals differently in terms of disruption and severity.

When extending the exposure periods, the effect of conflict on EVD cases was almost eliminated by 10 weeks in the DRC, while in Guinea, the risk of EVD cases decreased to 4 weeks and then increased by 10 weeks. However, it appears that extending the lag periods may have been diluting the effect of conflict on EVD cases, as changing the lag periods to a single week made all the lag periods for both countries more significant and impactful. These results track with previous research which found synchronicity and correlation between the reporting of EVD cases and conflict in the DRC and suggest that the effect of conflict may be long-lasting [11,19].

The results show that conflict is still highly influential multiple months following the event, and in some instances, conflict had a greater and more significant effect on EVD with time. A potential explanation may be due to the disruption caused by the conflict event having a cumulative effect. Previous findings suggest that conflict lengthens the EVD treatment process, e.g., longer times to detection, isolation, treatment and vaccination, creating a lasting effect and prolonging outbreaks [18]. Therefore, time would be needed for EVD cases to increase and be reported, after elevated transmission.

There are several limitations to the datasets used here, the first being the assumption that if EVD or conflict occurs, it is correctly identified (e.g., confirmed and suspected EVD cases were included) and reported on the correct day with no delay, where the transmission event/symptom onset occurred. These assumptions are unlikely, as EVD cases may be missed if cases never seek formal medical assistance or are in hard-to-reach areas. Furthermore, the categorisation of conflict events is subjective (category definitions were included in Additional file 1) and several categories are ambiguous. Data availability was limited and sub-setting the data (by sub-national unit or conflict sub-event type) led to small sample sizes in some instances, resulting in wide uncertainty, statistical insignificance or the models not being able to fit, particularly in Guinea. However, generally the findings were replicated in both countries, in terms of sub-event type, the effects of lag and geographic distribution of risk.

The methodology accounts for confounders by fixing them across the administrative unit. It is not as effective across multiple administrative units. Sub-national units are different geographical sizes, with different population sizes and risk factors. Therefore, comparing different regions/territories can create misleading results. Additionally, the use of vaccination during these more recent outbreaks [45,46], makes comparison to previous work challenging. While data on Ebola vaccination are available, the use of ring vaccination makes it difficult to incorporate vaccination data into the model used here (as a binary outcome) [38,46]. For example, the sub-national region with the largest IRR here was Équateur, part of the DRC which has received relatively little vaccination, compared to the eastern provinces [38].

### Conclusions

The research presented here highlights the increased risk that conflict has had on EVD cases in the DRC and Guinea in recent resurgences. The risks of conflict on EVD was spatially heterogenous and appears to last for several months and may be longer-lasting. Protests and riot-related conflict event types were highly influential, potentially being more disruptive to more people, compared to more violent forms of conflict e.g., battles. More research to understand how best to quantify conflict severity and intensity could provide further evidence for these mechanisms.

Extra vigilance is needed following protest- and riot-related conflict events for EVD transmission. Continued efforts are needed to try and account for the long-term effect of conflict on EVD and other infectious diseases. Effective public health messaging and scaling up testing in urban areas may help to mitigate these risks, including information on how to reduce transmission in dense urban areas and early clinical symptoms to be aware of. Ebola treatment centres should be prepared for an increase in cases following conflict events for multiple months, with sufficient vaccination stocks made available to areas most at risk.

## Data Availability

The datasets analysed during this study are all publicly available and references throughout the manuscript. Further code related to the methods and data visualization here are available at: https://github.com/GinaCharnley/ebola_conflict.

## List of abbreviations

ACLED: Armed Conflict Location & Event Data Project
CI: Confidence Interval
DRC: Democratic Republic of the Congo
EVD: Ebola Virus Disease
GUI: Guinea
HEMA: Humanitarian Emergency Response Africa
IRR: Incidence Rate Ratio
MoH: Ministry of Health
NGOs: Non-Governmental Organisations
SCCS: Self-Controlled Case Series
WASH: Water, Sanitation and Hygiene

## Declarations

### Ethics approval and consent to participate

Although the work presented here uses human data, all the data used are from public available data sources, which are none identifiable and therefore ethical approval was waived for this analysis.

### Consent for publication

Not applicable

### Authors’ contributions

GECC conceptualised and designed the study, collated the data sources, analysed the data, wrote the manuscript and incorporated feedback. IK contributed to interpreting the findings, provided expertise on conflicts and global health and revised several drafts of the manuscript. NG provided statistical expertise on the methodology, assisted in writing the manuscript, analysing the data and revised several drafts. EBM contributed to interpreting the findings, provided expertise on conflicts and Ebola in the DRC and revised several drafts of the manuscript. KAMG co-conceptualised and designed the study, provided data sources, provided expertise on epidemiological methods, contributed to interpreting the findings and revised several drafts of the manuscript. All authors read and approved the final manuscript.

## Acknowledgements

We would like to acknowledge and thank those working in the field who collected and collated the data sources used here, namely the Ministère de la Santé Publique de la République Démocratique du Congo and Ministère de la Santé et de l’Hygiène Publique de Guinée.

Additionally, we would like to thank Sophie Meakin for collating the situation reports used in some of the DRC data used here.

