## Additional file 1 for "Evaluating the Risk of Conflict on Recent Ebola Outbreaks in Guinea and the Democratic Republic of the Congo"

**Additional file 1**
Full definitions of conflict event types, defined by ACLED

ACLED data collect information on six types of events, both violent and non-violent, that constitute political disorder. These include:

1. Battles: Violent interactions between two organized armed groups;
2. Explosions/Remote violence: An event involving one side using remote weapons (e.g. artillery). These events can be against other armed actors, or used against civilians;
3. Violence against civilians: Violent events where an organized armed group deliberately inflicts violence upon unarmed non-combatants;
4. Protests: Public demonstrations in which the participants are not violent;
5. Riots: Violent events where demonstrators or mobs engage in destructive acts against property and/or disorganized acts of violence against people;
6. Strategic developments: Strategically important instances of non-violent activity by conflict actors and other agents within the context of conflict or broader political disorder. These can include recruitment drives, incidents of looting, and arrests are some examples of what may be included under this event type. Note that strategic developments are coded differently from other event types, and hence users must remember that they should be used differently from other event types in analysis.

Within these broad event categories, ACLED codes 25 sub-event types that classify different actions. Sub-event types allow for deeper analysis of events by “type”.

Source: https://acleddata.com/resources/quick-guide-to-acled-data/#s7

Further information and full documentation is available at: https://acleddata.com/acleddatanew/wp-content/uploads/dlm_uploads/2023/05/ACLED-Codebook-2023.pdf
