## Additional file 2 for "Evaluating the Risk of Conflict on Recent Ebola Outbreaks in Guinea and the Democratic Republic of the Congo"

**Additional file 2**
The layout of the pseudo-dataset dataframe fitted to the model.

The example shown here is for the Democratic Republic of Congo at exposure end point 1 (the week the conflict was reported). Each exposure and event (in the same administrative unit) are allocated a unique number (indiv). exday is the conflict (binary outcome = exgr) and eventday (binary outcome = event) is the EVD case. The start, end and interval refer to the exposure periods and the lengths between them.

Filter

| | indiv | exday | eventday | start | end | event | exgr | interval | loginterval |
|---|---|---|---|---|---|---|---|---|---|
| 1 | 1 | 2 | 79 | 1 | 2 | 0 | 0 | 1 | 0.000000 |
| 2 | 1 | 2 | 79 | 2 | 3 | 0 | 1 | 1 | 0.000000 |
| 3 | 1 | 2 | 79 | 3 | 183 | 1 | 0 | 180 | 5.192957 |
| 4 | 2 | 2 | 80 | 1 | 2 | 0 | 0 | 1 | 0.000000 |
| 5 | 2 | 2 | 80 | 2 | 3 | 0 | 1 | 1 | 0.000000 |
| 6 | 2 | 2 | 80 | 3 | 183 | 1 | 0 | 180 | 5.192957 |
| 7 | 3 | 2 | 81 | 1 | 2 | 0 | 0 | 1 | 0.000000 |
| 8 | 3 | 2 | 81 | 2 | 3 | 0 | 1 | 1 | 0.000000 |
| 9 | 3 | 2 | 81 | 3 | 183 | 1 | 0 | 180 | 5.192957 |
| 10 | 4 | 2 | 82 | 1 | 2 | 0 | 0 | 1 | 0.000000 |
| 11 | 4 | 2 | 82 | 2 | 3 | 0 | 1 | 1 | 0.000000 |
| 12 | 4 | 2 | 82 | 3 | 183 | 1 | 0 | 180 | 5.192957 |
| 13 | 5 | 2 | 83 | 1 | 2 | 0 | 0 | 1 | 0.000000 |
| 14 | 5 | 2 | 83 | 2 | 3 | 0 | 1 | 1 | 0.000000 |
| 15 | 5 | 2 | 83 | 3 | 183 | 1 | 0 | 180 | 5.192957 |
| 16 | 6 | 2 | 84 | 1 | 2 | 0 | 0 | 1 | 0.000000 |
| 17 | 6 | 2 | 84 | 2 | 3 | 0 | 1 | 1 | 0.000000 |
| 18 | 6 | 2 | 84 | 3 | 183 | 1 | 0 | 180 | 5.192957 |
| 19 | 7 | 2 | 85 | 1 | 2 | 0 | 0 | 1 | 0.000000 |
| 20 | 7 | 2 | 85 | 2 | 3 | 0 | 1 | 1 | 0.000000 |
| 21 | 7 | 2 | 85 | 3 | 183 | 1 | 0 | 180 | 5.192957 |

The data was fit to the model as follows: clogit(event ~ exgr + strata(indiv) + offset(loginterval), data = datLong). The data set up follows the work of Heather Whittaker, further code and examples are available at: http://stats-www.open.ac.uk/sccs/r.htm. The data are based on the examples related to multiple risk periods. The aim is to evaluate the likelihood of event = 1 and exgr = 1, vs event = 1 and exgr = 0. A pre and post exposure period are included to account for the possibility that the event could increase or decrease the probability of an exposure and because exposures can occur after the event. The interval is set up as an offset to account for that fact that a longer interval would increase the chances of the event occurring within it, not because the exposure increased the event but because there was a greater period of time for it to occur by chance.

Additional explanations of these assumptions are available at:
1. Petersen I, et al. Self controlled case series methods: an alternative to standard epidemiological study designs. BMJ 2016;354.
2. Farrington CP, et al. Case series analysis for censored, perturbed, or curtailed post-event exposures. Biostatistics 2009;10(1):3-16.
