## Additional file 3 for "Evaluating the Risk of Conflict on Recent Ebola Outbreaks in Guinea and the Democratic Republic of the Congo"

**Additional file 3**
Results of conflict event type on EVD cases in the DRC and Guinea

Battles, riots and protests had the strongest effect on EVD cases across the DRC and Guinea (Fig. S1), however in Guinea, battles, protests, strategic developments and violence against civilians were not significantly associated with EVD cases here. IRR values showed an increased risk of certain types of conflict, compared to the national levels. The strongest effect included battles and riots in Guinea at 3.45 (0.80-14.8 95%CI) and 2.57 (1.31-5.02 95%CI) times increased risk of EVD cases, while in the DRC, protests, violence against civilians and battles had the greatest effect on EVD, with IRR values of 2.10 (1.83-2.40 95%CI), 1.79 (1.63-1.95 95%CI) and 1.78 (1.62-1.94 95%CI), respectively.

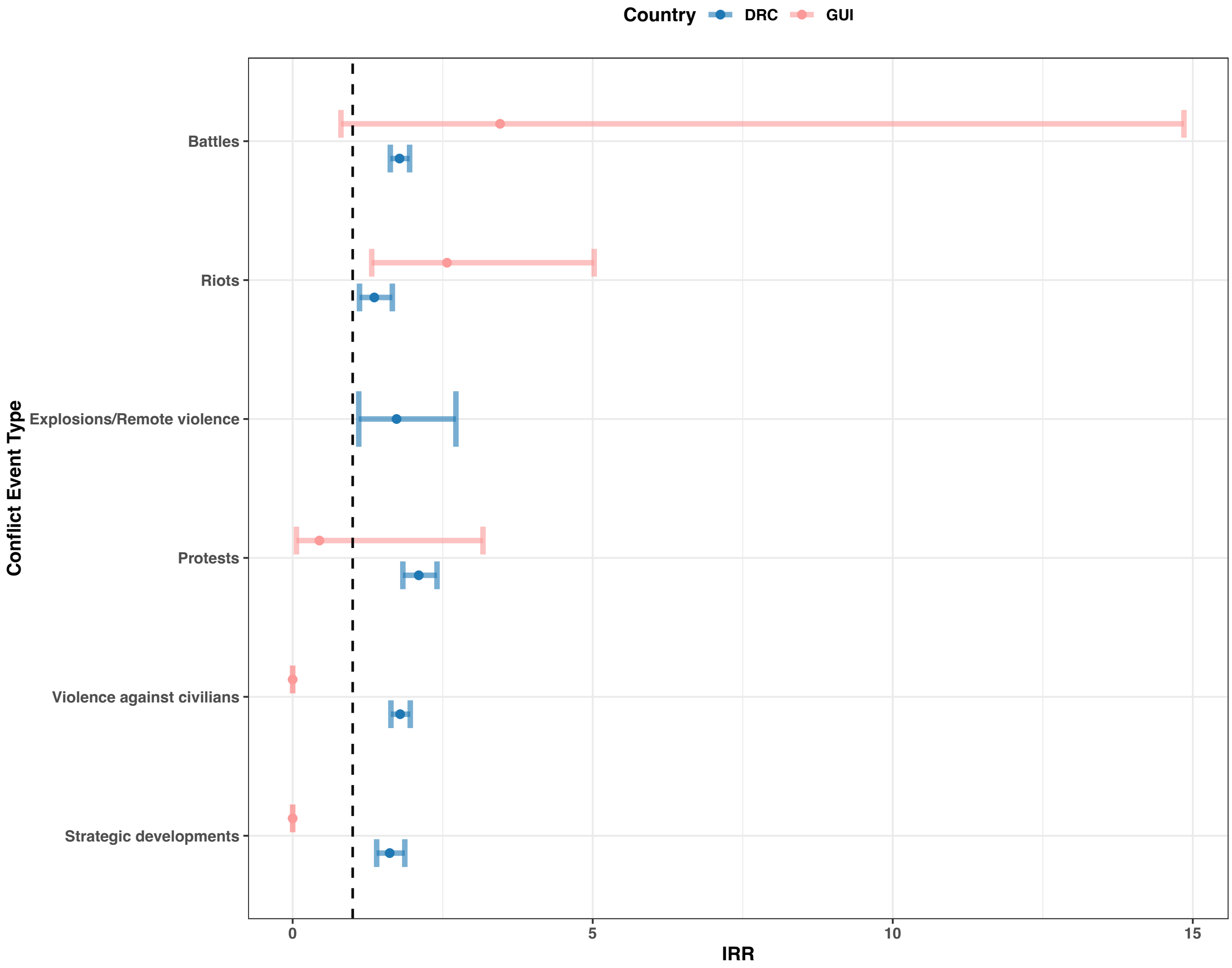


**Fig. S1** Incidence rate ratio (IRR) for the effect of conflict event exposure on EVD cases for the Democratic Republic of Congo (DRC) and Guinea (GUI) during the week the conflict was reported by conflict event type.
