## Additional file 4 for "Evaluating the Risk of Conflict on Recent Ebola Outbreaks in Guinea and the Democratic Republic of the Congo"

Proximity of battles, protests and riots to major cities in Guinea and the DRC

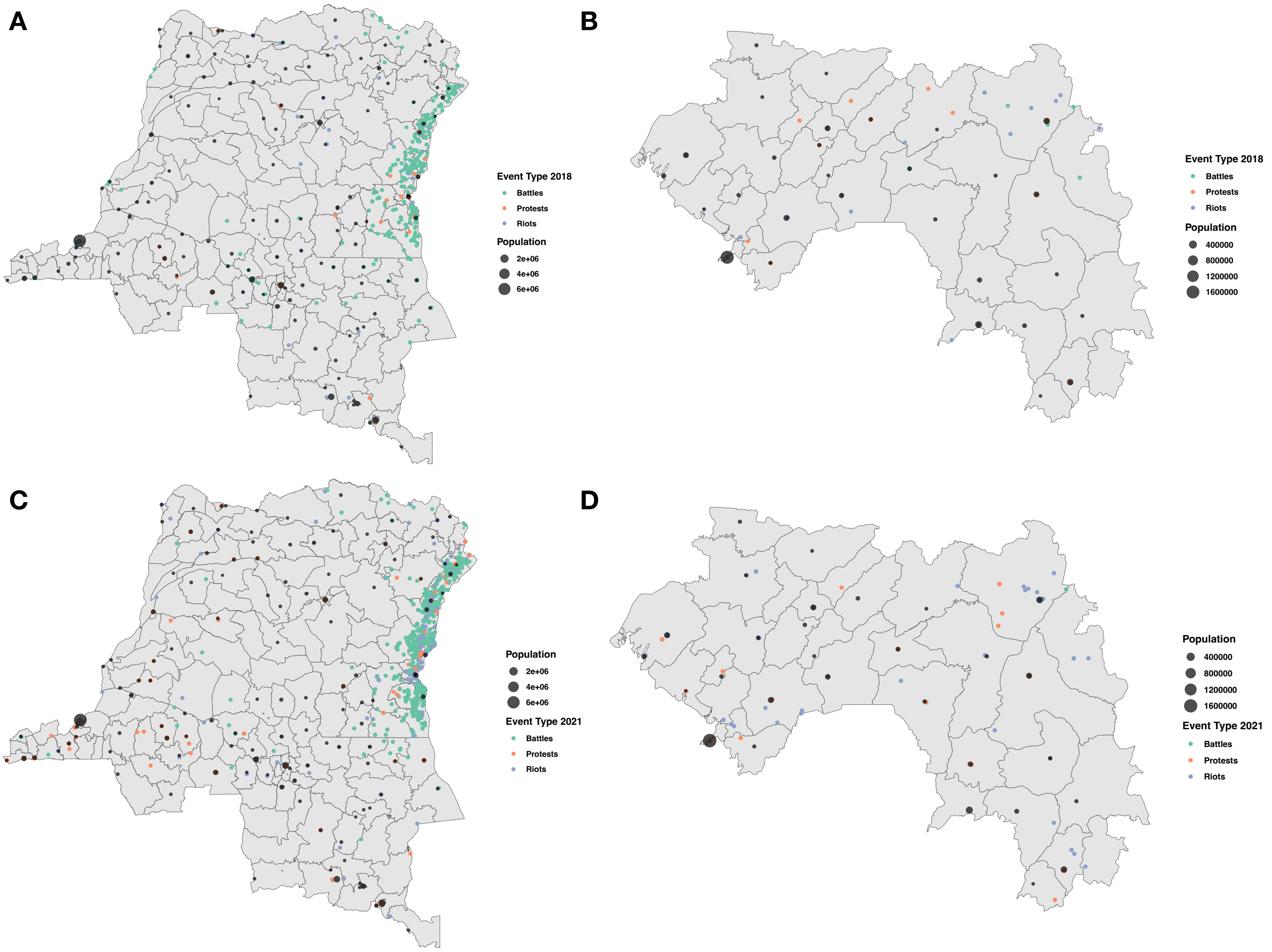


**Fig.S2** The location, by longitude and latitude, of major cities (point size = population size) and conflict event for **A**, the Democratic Republic of Congo in 2018, **B**, Guinea in 2018, **C**, the Democratic Republic of Congo in 2021 and **D**, Guinea in 2021.
